# Adaptive trials in stroke: Current use & future directions

**DOI:** 10.1101/2024.04.04.24305236

**Authors:** Kathryn S Hayward, Emily J Dalton, Bruce CV Campbell, Pooja Khatri, Sean P Dukelow, Hannah Johns, Silke Walter, Vignan Yogendrakumar, Jeyaraj Pandian, Simona Sacco, Julie Bernhardt, Mark Parsons, Jeffrey L Saver, Leonid Churilov

## Abstract

Inclusion of adaptive design features in a clinical trial provides pre-planned flexibility to dynamically modify a trial during its conduct, while preserving validity and integrity. Adaptive trials are needed to accelerate the conduct of more efficient, informative, and ethical clinical research in the field of neurology as compared to traditional fixed designs. Stroke is a natural candidate for adoption of these innovative approaches to trial design. This Research Methods in Neurology paper is informed by scoping review that identified 45 completed and ongoing adaptive clinical trials in stroke that were appraised: 14 trials had published results with or without a published protocol, 15 trials had a published protocol, and 16 trials were registered only. Treatments spanned acute (n=28), rehabilitation (n=8), prevention (n=8), and rehabilitation and prevention (n=1) domains. A subsample of these trials were selected to illustrate the utility of adaptive design features and discuss why each adaptive feature(s) were incorporated in the design to best achieve the aim, whether each individual feature was used and if it resulted in expected efficiencies, and any learnings during preparation, conduct or reporting. We then discuss the operational, ethical, and regulatory considerations that warrant careful consideration during adaptive trial planning and reflect on the workforce readiness to deliver adaptive trials in practice. We conclude that adaptive trials can be designed, funded, conducted, and published for a wide range of research questions and offer future directions to support adoption of adaptive trial designs in stroke and neurological research more broadly.

## Introduction

Methodologists have long signalled that inclusion of adaptive design features in clinical trials can drive more efficient and ethical research across clinical trial phases as compared to traditional fixed designs. An adaptive clinical trial offers pre-planned opportunities to use accumulating data to modify design aspects during trial conduct while preserving validity and integrity^1^. Adaptations are implemented for the purpose of both maximising statistical (and at times operational) efficiency and achieving better outcomes for participants and future patients^2^. Such goals can be achieved through a variety of adaptive features used individually or in combination (**Table 1**), with the most complex option being an adaptive platform trial. Common features include adapting the sample size to gain sufficient power based on observed outcomes, removing treatments that are less effective, adapting randomisation ratios in response to treatment outcomes, and enriching recruitment to specific subgroups that appear most likely to benefit. Enacting pre-planned adaptive features due to futility may translate to fewer patients allocated to ineffective treatments, which presents a possible cost-saving.

**Table 1:** Glossary of common adaptive design features synthesised from guidance documents^1, 2, 5^.

|  |
| --- |
| **Adaptations to the timing for stopping a trial** (also referred to as group sequential): Allows for one or more prospectively planned interim analyses of data with prespecified criteria for stopping the trial. Criteria for stopping may be for early efficacy, safety, or insufficient evidence of efficacy, which is often called stopping for futility. |
| **Adaptations to sample size:** Allows adaptive modification to the sample size based on interim estimates that uses treatment assignment information or comparative interim results. |
| **Adaptations to patient population:** Allows adaptive modifications to the patient population and often involves both (a) modification of design features, such as the enrolled population and the population evaluated in the primary analysis, based on comparative interim results; and (b) hypothesis tests in multiple populations, such as a targeted subpopulation and the overall population. |
| **Adaptations to patient allocation based on comparative baseline characteristic data:** Covariate-adaptive randomisation (CAR) is adaptive modification to treatment assignment depending on subject's baseline characteristics, and the treatment assignments of previously enrolled subjects to reduce the covariate imbalance in treatment arms. |
| **Adaptations to patient allocation based on comparative outcome data:** Response-adaptive randomisation (RAR) is adaptive modification to treatment assignment based on accumulating outcome data of subjects previously enrolled. |
| **Adaptations to treatment arm selection:** Allows adaptive modification to the treatment arms based on comparative interim result which may see arms added or terminated. |
| **Adaptations to endpoint selection:** Allows adaptive modification of the primary endpoint based on comparative interim results. |
| **Adaptive platform trial:** Allows for the study of multiple interventions in a single disease (or condition) in a perpetual manner, with interventions allowed to enter or leave the platform based on a decision algorithm. This design is based on multiple arms studied over multiple stages (MAMS) with a common control group(s). Such designs are often driven by within trial adaptations (e.g., adaptations to patient allocation based on comparative outcome data). |

Inclusion of adaptive features in clinical trials has been growing since the early 2000s^3, 4^. Several regulatory agencies have published guidance statements^5, 6^. In 2020 the Adaptive designs CONSORT Extension (ACE) statement was published^1^. Such works have motivated funding bodies, researchers, and networks to consider the value of clinical trial designs that include adaptive features to deliver their research agenda^7, 8^. While historically the leading area of adaptive trial application was oncology^3, 4^, accelerated implementation was observed in the context of the COVID-19 pandemic^9^. Despite adaptive trials being proposed as a possible solution to improve clinical trial conduct^10^, adoption in many conditions^3, 4^ including stroke is not well documented.

Clinical trials of treatments for people with stroke are well suited to adaptive designs^11^. There are many treatments across acute, rehabilitation, and prevention domains of the heterogeneous disease of stroke^11–14^ that need to be efficiently screened and tested for clinical efficacy. This highlights the urgency to advance the use of adaptive trial designs to identify early futility and eliminate ineffective treatments during trial conduct. Such impacts can prevent delays in testing alternative treatments, whilst improving resource efficiency and minimising how many patients are exposed to ineffective (and potentially harmful) treatments^11^. As accumulated data inform adaptations, the very nature of an adaptive trial depends on the outcome of interest being observed precisely over a period that is substantially shorter than the overall trial duration in order to inform pre-planned adaptations. Primary outcomes for stroke trials can be measured as early as within the first 24 hours (e.g., early neurological recovery or imaging outcomes^15, 16^) to 90 days post-stroke (e.g., modified Rankin Scale in acute ischaemic stroke^17^, and rehabilitation^18^) and may even extend out to 1 year and beyond (e.g., intracerebral haemorrhage^19^, or composite outcomes in long-term prevention^20–22^ trials). Some early outcomes (e.g., reperfusion within 24 hour after ischaemic stroke) are also highly prognostic of longer-term functional outcome (e.g., modified Rankin Scale at 90 days)^23^. Collectively, this positions stroke as a natural candidate for embracing adaptive clinical trial designs.

This paper uses a scoping review methodology to identify completed and ongoing adaptive clinical stroke trials (acute, rehabilitation, prevention) to inform real-world case examples that illustrate the utility of adaptive design features in practice. We subsequently consider the strengths and limitations of adaptive designs and workforce readiness to deliver adaptive trials (including the role of stroke organisations and trial networks to support their uptake). We conclude with future directions, for stroke and neurology broadly, to support adoption of adaptive trial designs to address important clinical research questions.

### Scoping review of adaptive clinical trials in stroke

Forty-five trials were identified from a scoping review of clinical trial registries and Pubmed (See Supplementals A-B). Fourteen adaptive trials had published results with or without a published protocol, 15 had a published protocol and 16 had registration only. Details of included trial characteristics, design, and adaptive features used and enacted is provided in Supplementals B-E. Treatments spanned acute (n=28), rehabilitation (n=8), prevention (n=8), and rehabilitation and prevention (n=1). The most common adaptive features were group sequential (n=29), and sample size reestimation (n=17). High use of group sequential is consistent with other observations^24^ and may relate to its broad definition (Table 1). Twenty-six trials included one adaptive feature, 12 included two and six included three or more. Most trials were investigator initiated (n=26 investigator-initiated only, n=12 investigator-initiated, industry-supported) with a high-income country-specific national agency being the primary funding source e.g., National Institutes of Health. Only three trials included sites in upper middle-income countries and four included sites in lower-middle-income country.

### Utility of adaptive features in stroke trials

Each adaptive feature (Table 1) was linked to a real-world stroke trial (selected from Supplemental B-E) to illustrate utility in written and visual format (Figures 1-5). Trials presented span clinical domains (acute/rehabilitation/prevention), trials phases and stroke aetiologies. One adaptive platform trial in the planning stage is also presented. Each trial discussion focuses on why the adaptive feature(s) were incorporated in the design to best achieve the aim, reflection on whether each individual feature was used and whether it resulted in expected efficiencies, and any learnings during preparation, conduct or reporting.

**Figure 1:**
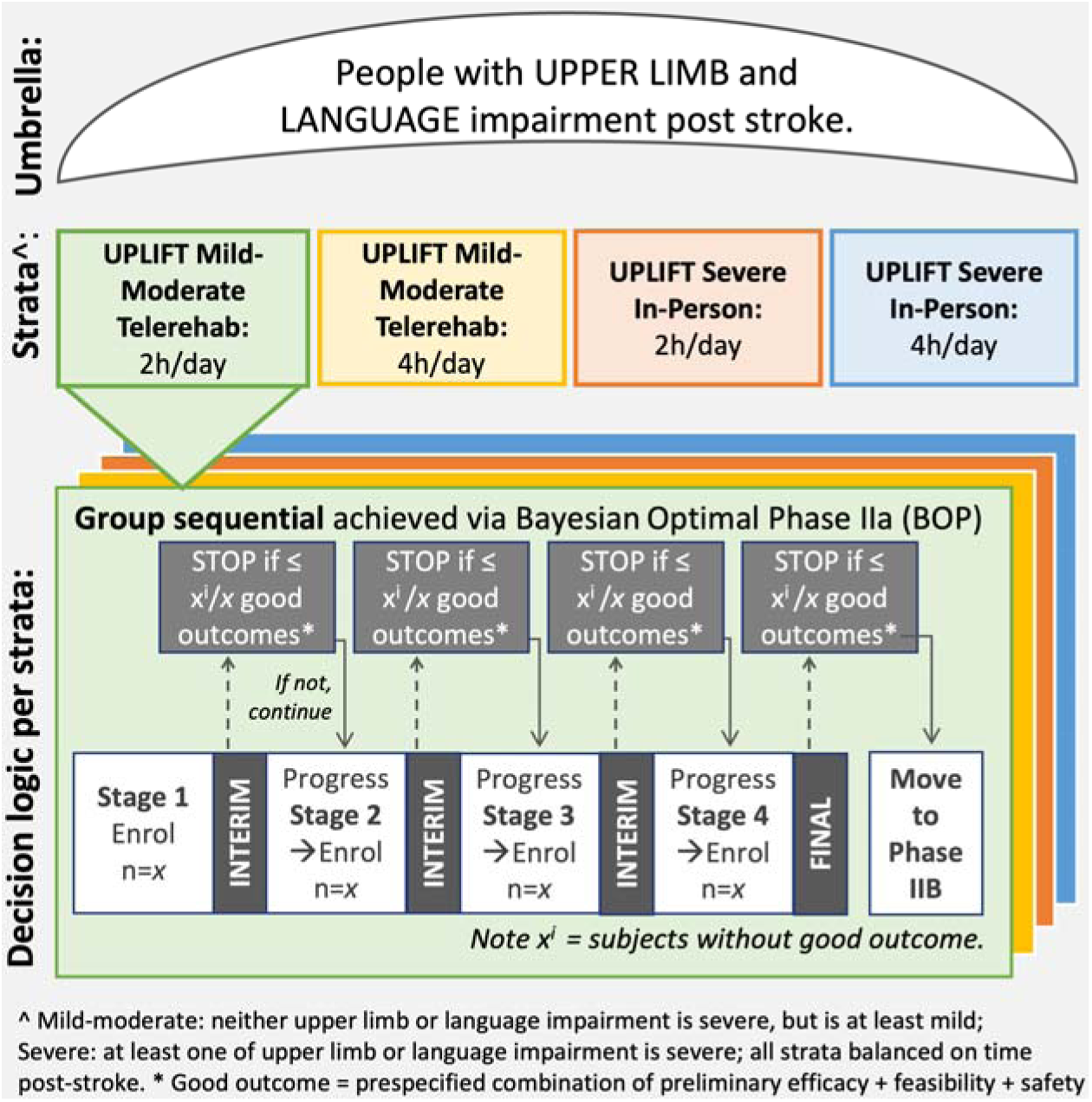
Visual representation of the adaptive feature of group sequential testing achieved via a Bayesian Optimal Phase II (BOP) design in the UPLIFT trial.

#### Adaptations to the timing for stopping a trial

The SELECT-2^25^ and TENSION^26^ acute trials tested the efficacy of endovascular thrombectomy in patients with extensive ischaemic injury. As there were potential safety and futility concerns in the most severely affected patients, their chosen designs included group sequential testing with strict z-score boundaries to reduce the risk of stopping early for false positive results. Due to early evidence of efficacy, the SELECT-2 stopped after the second interim analysis results, and the TENSION trial stopped after the first interim analysis results were available. Due to ongoing recruitment while the interim analysis outcomes were accrued, the final sample size was larger, and this was tested at the conventional significance level in the final analysis. On completion, both trials demonstrated that among patients with large ischaemic strokes, endovascular thrombectomy resulted in better functional outcomes than medical care. These early and positive findings resulted in cost, resource and time savings for the trial and future participants and led to rapid inclusion into clinical practice guidelines e.g., SELECT-2 included in the living Australian Stroke Clinical Practice Guidelines within 5-months of publication. It must be noted though that in SELECT-2 only, endovascular thrombectomy was associated with early neurologic worsening and procedural complications.

The UPLIFT trial (integrated UPper limb and Language Impairment and Functional Training, ACTRN12622000373774) is an ongoing rehabilitation trial that includes group sequential testing. This trial aims to efficiently develop evidence for a new model of stroke rehabilitation, during community living. An umbrella design with four simultaneous Bayesian Optimal Phase IIa (BOP)^27^ strata was adopted. In this design, the outcomes from the four BOP strata are not compared to each other but rather to a pre-specified objective criterion^28^. The overall goal is to select the dose(s) of the UPLIFT intervention presenting with sufficient promise, probed through group sequential testing (three interim analyses, one final decision analysis per strata). This offers a flexible approach to identify promising individual interventions by screening multiple doses under a single trial infrastructure. The adopted compositive binary primary good outcome (inclusive of signal of efficacy, safety, and feasibility) is used to determine if a specific individual UPLIFT intervention is stopped or continued at each interim analysis point. The field of stroke recovery is yet to deliver practice changing interventions like thrombolysis or thrombectomy. There is a time imperative to identify futility early for novel interventions, which was afforded by a group sequential testing.

During trial set-up, ethical approval was efficient (<30 days from submission to approval), which was afforded by clear reporting of the adaptive design due to statistical input and oversight, and use of a design overview figure **(Figure 1)**. However, there were challenges associated with trial registration as the process at the time was not customised to cover for umbrella designs with adaptive features.

#### Adaptations to sample size

The EXTEND-IA TNK trial^29^ compared two different clot-dissolving medicines (alteplase and tenecteplase) prior to EVT in a phase II non-inferiority design using reperfusion at the initial angiogram as the primary outcome. Given the uncertainty around the expected effect size, an *adaptive sample size reestimation* was employed using the Mehta and Pocock promising zone method^30^ **(Figure 2)**. This has the advantage of being fully prespecified and reestimates the sample size within a specified range, based on the conditional power observed at the time of sample size reestimation. The choice of when to perform the interim sample size reestimation is a balance between greater precision with larger participant numbers and sufficient time to accumulate evidence regarding the primary outcome, accounting for recruitment rate, prior to reaching the minimum sample size. In EXTEND-IA TNK, the primary outcome was collected within hours of enrolment. This short period over which to observe the outcome allowed the reestimation to occur closer to the originally planned full sample size than it would have been had an outcome been collected at 3 months. The prespecified mechanism of the Mehta and Pocock method does not impose alpha-spending penalties for multiplicity of testing and uses the threshold of p=0.05 for the final analysis. It is important to note that the intent here is quite different to that used by group sequential testing which imposes a stringent (e.g., p<0.001) early stopping boundary to demonstrate early efficacy. In this trial, the estimated sample size was set at a minimum of 120 with a potential increase up to the maximum of 276 patients. The prespecified sample size reestimation occurred at 100 patients and established a final sample size of 202 patients. On completion, it was demonstrated that tenecteplase before thrombectomy was associated with a higher incidence of reperfusion and better functional outcome than alteplase among patients with ischaemic stroke treated within 4.5h after symptom onset.

**Figure 2:**
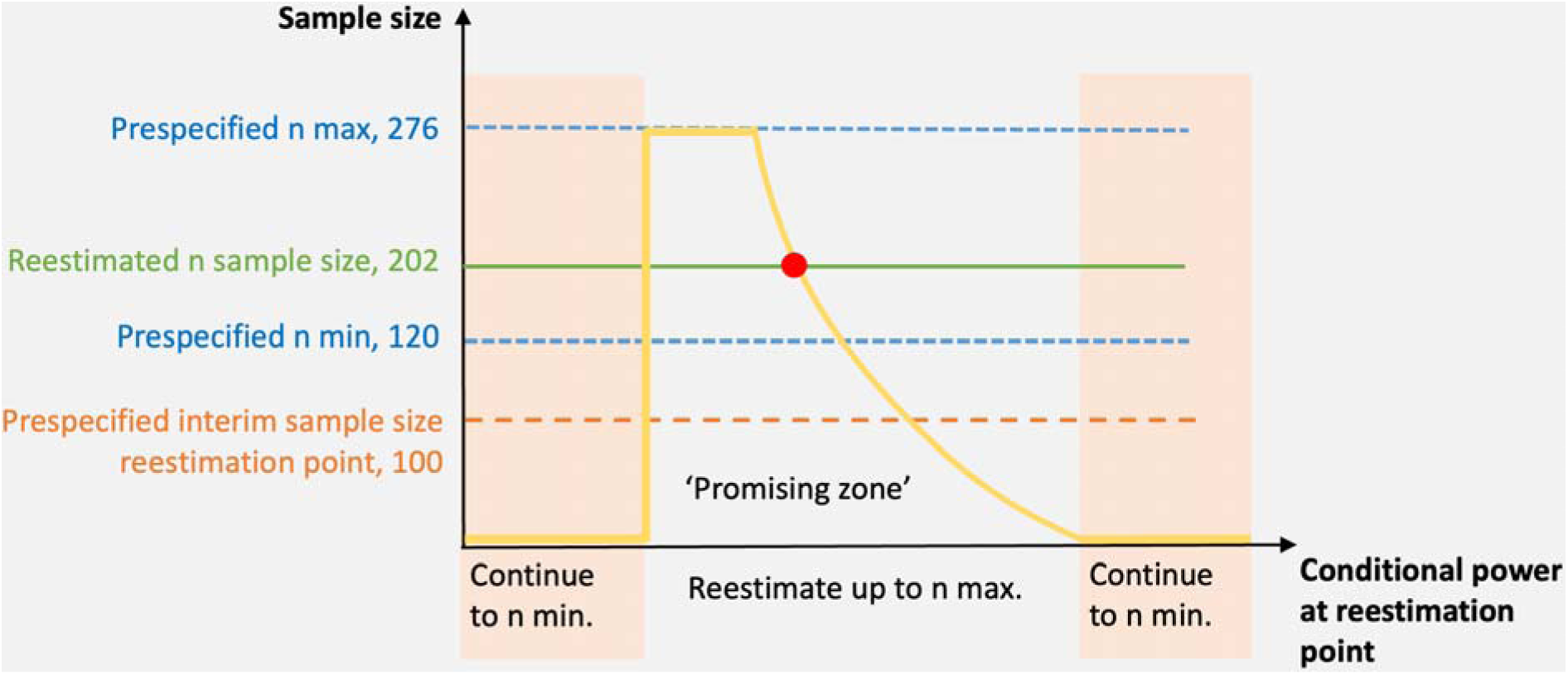
Visual representation of the adaptive feature of sample size reestimation achieved using the Mehta and Pocock promising zone method in the EXTEND-IA TNK trial.

#### Adaptations to patient population

ENRICH is a randomised trial^31^ of minimally invasive surgery for intracerebral haemorrhage (ICH) that has been presented in abstract form^32^. The trial used a Bayesian design with adaptive enrichment. Scheduled interim analyses, beginning after 150 patients (and then at 175, 200, 225, 250 and 275, with the trial maximum sample of 300 patients) had been enrolled were used to guide study enrichment based on haemorrhage location. A visual representation of how adaptive enrichment works is provided in **Figure 3**. At interim analysis (adaptive feature of group sequential testing), the anterior basal ganglia stratum was closed early due to futility according to prespecified thresholds for enrichment. The final trial results were positive, driven by the benefit in enriched lobar ICH, with neutral effect in the anterior basal ganglia group. ENRICH is the first trial to enact adaptive enrichment. The SELECT-2^25^, DEFUSE 3^33^ and DAWN^34^ trials all pre-specified the potential for adaptive enrichment to allow for sequential exclusion of subpopulations that were likely to have the worst prognosis and enrich the remaining trial population, should the overall interim result cross a safety or futility boundary. However, none of these three trials needed to enact adaptive enrichment.

**Figure 3:**
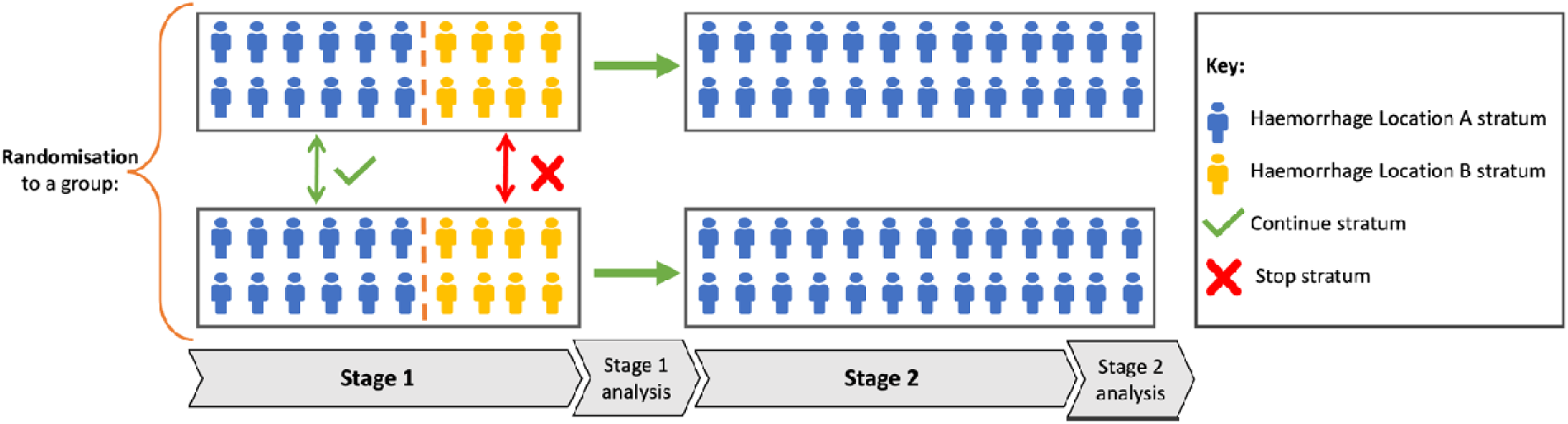
Visual representation of adaptations to the patient population, in this example, based on stratum of haemorrhage location.

#### Adaptations to patient allocation based on comparative baseline characteristic data

The TELEREHAB-2 trial^35^ was designed to determine whether treatment targeting arm movement delivered via a home-based telerehabilitation system had comparable efficacy with dose-matched, intensity-matched therapy delivered in a traditional in-clinic setting, and to examine whether this system has comparable efficacy for providing stroke education. Baseline characteristics were selected as covariates, based on established evidence on potential confounding, in an attempt to prevent serious imbalances across the two treatment groups. The covariates used in this trial were time post-stroke, severity of impairment, age, enrolment site, and stroke subtype were balanced across the two treatment groups. **Figure 4** provides a visual representation of this adaptative feature in the context of the AVERT-Dose trial.

**Figure 4:**
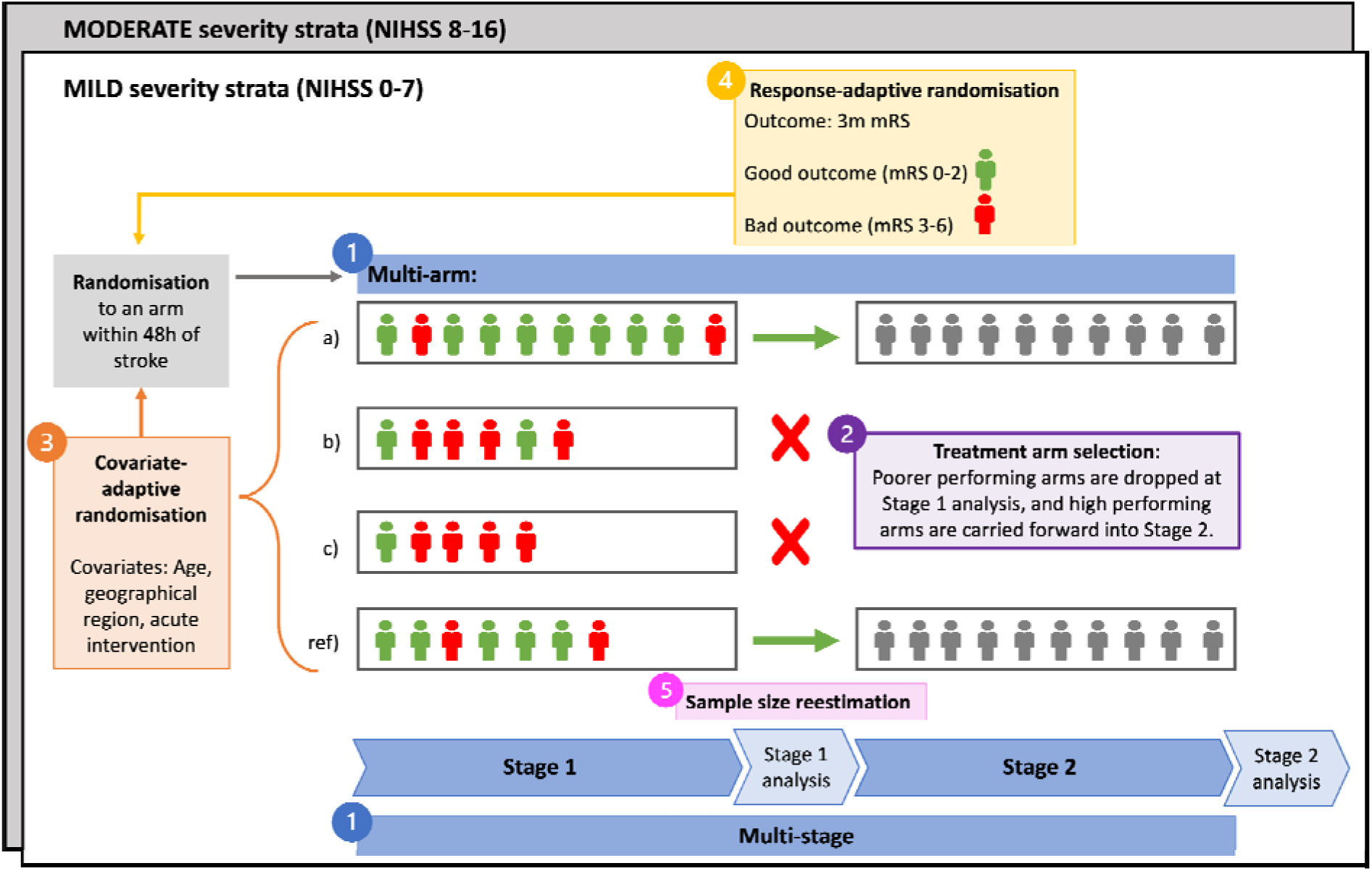
Visual representation of the five adaptive features (numerically numbered) of (1) multi-arm, multi-stage, (2) treatment arm selection, (3) covariate-adaptive randomisation, (4) response-adaptive randomisation and (5) sample size reestimation included in the AVERT-Dose trial.

#### Adaptations to patient allocation based on comparative outcome data

MOST^31^ is an ongoing multi-arm trial of stroke thrombolysis to determine the safety and efficacy of intravenous therapy with argatroban or eptifibatide as compared with placebo in acute ischemic stroke patients treated with intravenous recombinant tissue plasminogen activator within 3 hours of symptom onset.

Interim analyses were planned to occur after 500, 700, and 900 subjects were randomised. Using a Bayesian approach, the predictive probability of a successful final analysis was calculated based on different assumptions about the remaining subjects to be enrolled. When n=500, one (or both) experimental arm(s) may be stopped for futility if there is less than 20% probability of demonstrating benefit compared to the control arm in either experimental intervention and the trial were to continue to n=700. Next, one (or both) experimental arm(s) may be stopped for futility when n=700 or n=900 if there is less than 5% probability of demonstrating benefit in either experimental arm if the trial were to continue. One (or both) arm(s) may be stopped early for efficacy after 700 or 900 subjects if an arm has an expected successful predictive probability of demonstrating superiority to control of at least 99%. This trial, reported at International Stroke Conference 2024, was stopped for early futility. **Figure 4** visually represents adaptations to patient allocation in the context of the AVERT-Dose trial.

#### Adaptations to treatment arm selection

The AVERT-DOSE trial (ACTRN12619000557134)^36^ is an ongoing rehabilitation trial that includes five adaptive features, including adaptation to treatment arm selection. This trial aims to define optimal early (commenced within 48 hour) mobility intervention regimens for ischaemic stroke patients of mild and moderate severity across seven countries (Australia, Malaysia, United Kingdom, Ireland, India, Brazil, Singapore). Ongoing uncertainty around the world about the safest and most effective early training approaches after stroke prompted this clinical trial^37^. The developed adaptive design is one of the most complex designs ongoing in stroke as it includes multi-arm, multi-stage with treatment selection, covariate-adjusted and response-adaptive randomisation, and sample size reestimation (**Figure 4**). Unlike the MOST trial, AVERT-Dose is performing response-adaptive randomisation within frequentist rather than Bayesian framework. Focusing on the adaptive feature of treatment selection, in Stage 1 (mild or moderate severity strata), 25% of patients will be randomised into the reference arm, while randomisation into three intervention arms is guided by the adaptive algorithm. At Stage 2, randomisation into the reference arm and the single selected intervention arm will be guided by the adaptive algorithm. Implementing this adaptive trial design enabled a wider variety of mobility regimes (three regimens per strata) to be tested than a traditional two-arm design, enabling more efficient testing than otherwise possible.

#### Adaptations to endpoint selection

The EVACUATE trial (NCT04434807) is an ongoing trial testing ultra-early minimally invasive evacuation of intracerebral haemorrhage (ICH). This has been proposed as a means to reduce ICH growth, secondary injury from mass effect, thrombin, and iron toxicity. The MISTIE-III trial^19^ provided proof of principle that effective ICH evacuation may reduce disability with a strong association between the proportion of ICH evacuation or the residual volume with functional outcome. This study also justified the use of the proportion of haematoma evacuated as an intermediate outcome (endpoint) in the EVACUATE seamless phase IIb/III two-arm, two-stage randomised trial (NCT04434807) with the primary outcome (endpoint) of mRS at 90 days (**Figure 5**). A two-arm, two-stage design is a specific case of a multi-arm, multi-stage adaptive design^11^ that capitalises on the principles of group sequential testing and complements it with additional useful adaptive features. A seamless IIb/III design is particularly advantageous in a relatively rare condition such as ICH, in that the phase IIb patients can also contribute to the phase III analysis of functional outcome. There are also logistical efficiencies in being able to proceed directly from phase IIb to III without stopping to re-submit ethics, obtain governance, and reinitiate sites. However, the use of an intermediate outcome to determine progression of the trial to the second stage (Phase III) is fully predicated on demonstrated prognostic properties of this outcome in relation to the primary outcome. In this trial, the two-arm, two-stage design was complemented by an adaptive sample size reestimation within the second stage using the Mehta and Pocock method described above^30^. The randomisation for this trial uses the covariate adaptive common scale minimum sufficient balance algorithm^38^ that improves the balance of key prognostic variables while maximizing the randomness of allocation.

**Figure 5:**
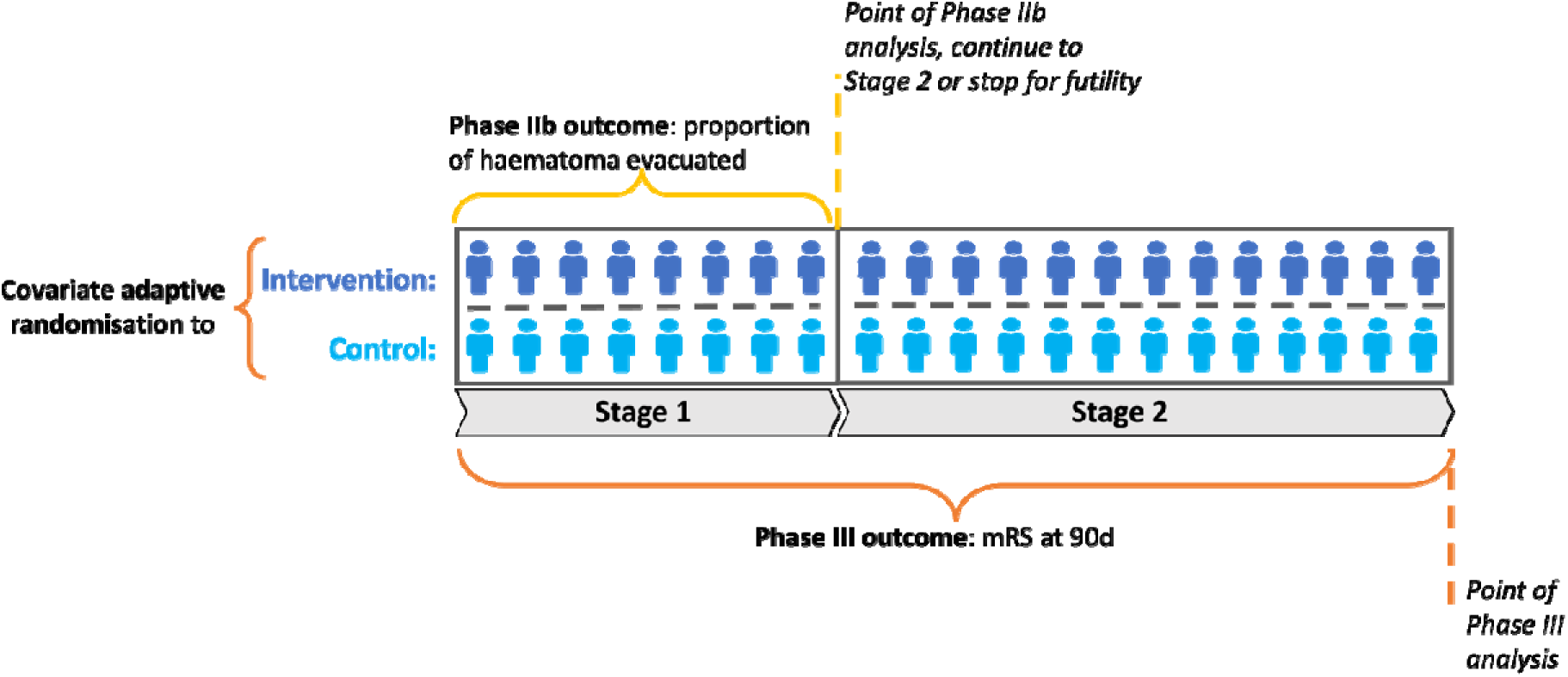
Visual representation of adaptations to the endpoint outcome selected at Phase IIb (haematoma evacuation) and Phase III (mRS at 90 days).

#### Planned adaptive platform trial

Recent pivotal trials have shown endovascular thrombectomy to have large treatment effects in highly selected subsets of patients with acute ischaemic stroke due to large vessel occlusions. These have raised questions about whether more broadly selected patients would benefit from endovascular therapy. Furthermore, despite large treatment effects, most endovascularly treated patients remain functionally disabled, raising many new treatment and management questions in urgent need of answers. Each of these questions would need large and expensive clinical trials if performed in a traditional manner, and major therapeutic advancements would be prohibitively expensive. Therefore, the StrokeNet Thrombectomy Endovascular Platform (STEP) has been designed within NIH StrokeNet to address these questions using statistical and operational efficiencies. STEP is designed to determine the optimal treatment strategies for patients with acute ischaemic stroke due to large or medium vessel arterial occlusions potentially amenable to endovascular therapy. The planning grant is nearing completion (NIH/NINDS OT2NS129366) and initial enrolment in the platform trial is anticipated in 2024.

STEP is a randomised, multi-factorial adaptive platform trial (NCT06289985) that aims to study which ischaemic stroke patients should be treated with endovascular therapy and how to optimise that care further, in terms of newer devices or techniques, adjunctive therapies, newer diagnostic strategies, and systems of care delivery^39^. The primary outcome is global functional outcome measured by the utility-weighted modified Rankin Scale assessed at 90 days. The platform will include 50 comprehensive stroke centres across the US and Canada. The sites will work under a master protocol that integrates appendices that address distinct interventions, or domains. Domain-specific modelling are integrated into one inferential statistical model, and the model allows for frequent adaptive analyses to assess the effects of distinct interventions.

The first domain, STEP Endovascular Thrombectomy Indication Expansion (STEP-EVT Indication Expansion, NCT06289985), will assess endovascular thrombectomy versus medical management in acute ischaemic stroke patient subgroups with low NIHSS and/or medium and distal vessel arterial occlusions. Patients will be concurrently randomised to other domains, as they become funded by the NIH and initiated within the platform, and the model will estimate the effects of other domains using that domain’s specific modelling. Adaptive conclusions for each domain will be triggered by the primary Bayesian model run at a prespecified frequency. Adaptive decisions for each domain may include response adaptive randomisation.

An adaptive platform trial, such as STEP, demonstrates how access to initial planning funds is critical to support the development of the platform and supporting infrastructure. Appropriate planning is important given adaptive platform trials can run for a long period of time. National funding agencies in USA, Australia, and UK have recently had targeted funding calls for adaptive platform trials. An important feature of a platform trial is a shared control group, which allows new treatments to be compared to a common control, direct comparison of effect sizes between treatments and reduces the likelihood that a patient will end up in the control group^9, 40^. However, use of a shared control does rely on the assumption that it remains equally acceptable to patients with time^9^. There are other considerations related to chronological bias including trends in outcome measures that may be caused by changes in standard of care, adding or dropping of participating centres, shifts in the patient population, systematic variations in participant responses to the control or changes in expectancy effects when a new treatment that is perceived as particularly promising enters or leaves the trials^9, 40^.

### Strengths and limitations of adaptive trials

Adaptive trial designs are heralded as a suite of tools to deliver more efficient trials. There are many aspects to consider whether this goal is achievable. Depending on the specific clinical research question, adaptive features (**Table 1**) may or may not be appropriate. The very nature of an adaptive trial depends on the outcome of interest being observed over a period that is substantially shorter than the overall trial duration in order to inform pre-planned adaptations. As such, adaptive trials may not be suitable when the outcome of interest is collected years later e.g. long follow-up in secondary prevention trials for stroke or myocardial infarction. Alternatively, there may be limited or no access to appropriate statistical design and analysis support. Adaptive trials may increase the practical complexity and in doing so, eliminate the theoretical efficiency gains^41^. **Table 2** provides a narrative synthesis of existing operational, ethical, scientific, and regulatory considerations. As this review is primarily for a clinical readership, discussion of the statistical considerations of adaptive designs^1, 24^ is beyond its scope.

**Table 2:** Narrative synthesis of operational, ethical, scientific, and regulatory considerations.

| Operational | Possible strengths provided | Possible limitations introduced |
| --- | --- | --- |
| Timely delivery of trial | Design, statistical and recruitment efficiencies can lead to shorter trial duration which may be seen as increased feasibility to deliver on time<sup>5</sup>. | Preplanning adaptive design modifications can require more effort at the design stage, leading to longer lead times between planning and starting the trial<sup>5</sup>. |
| Funding agencies | Adaptive trials can be viewed as novel and innovative, which can see them embraced. Examples of this include recent funding agency statements and calls from the Medical Research Future Fund in Australia<sup>7</sup>, National Institutes of Health in USA<sup>52</sup>, and National Institute of Health and Care Research in UK<sup>8</sup>. | Conservative decisions by funding agencies may limit development of an understanding of when adaptive trials are applicable, what they can (and cannot accomplish), what their practice implications are, and potential results expected<sup>24</sup>. Can be challenging to secure funding for some adaptive features e.g., for a seamless Phase IIb-III, some funding agencies may want to see Phase IIb results before providing Phase III funding. |
| Infrastructure | (electronic)Case Record Forms built may be used for more than one study e.g., master protocol studies of adaptive platform, basket, or umbrella<sup>9, 40</sup>. | Shared trial infrastructure must be hosted and maintained by some entity, which may change or evolve with time<sup>9</sup>. Infrastructure must be built to be flexible and scalable from the outset, which required considerable planning and cost investment<sup>53</sup>. |
| Costs | Median increase in cost compares favourably against the increase in efficiency that some adaptive designs can introduce e.g., group sequential<sup>54, 55</sup>. | Increased costs have been identified across all aspects of an adaptive trial, with the greatest increase in statistical and design related costs<sup>43</sup>. |
| Logistics | Introducing adaptive designs from early conceptualization of the trial is more effective than fitting an adaptive design at later stages of design<sup>56</sup>. Allowing sufficient time for the design process to evolve and garner input from clinicians, biostatisticians, adaptive design and lived experience experts is necessary<sup>56</sup>. | Logistical challenges can include limited access to resources for planning and technical requirements prior to funding submissions<sup>56</sup>, communicating protocol changes to all parties<sup>57</sup>, costing and forecasting budgets<sup>43</sup>, planning for product supply, access to high-quality interim data in a timely manner so that adaptive decision-making can be based on up-to-date and reliable results and the potential need for real-time Data and Safety Monitoring Board decisions<sup>5</sup>. |
| Recruitment | Enhanced targeting of recruitment to the 'right' participants can arise during trial conduct due to inclusion of some adaptive features. Features of adaptive randomisation or population enrichment have been viewed positively by enrolling participants<sup>58</sup>. | Cumbersome recruitment, identification, and consenting processes can be major barriers, especially at more novice trial centres<sup>59</sup>. This can limit adaptive trials to primarily occur at large, academic medical centres with existing research infrastructure<sup>59</sup>. |
| Staffing | Staff working on adaptive trials have acknowledged that they were | Increased demands on staff do exist, especially master protocol adaptive trials where |
|  | exposed to and gained experience in all stages of a trial in a much shorter window of time compared to more traditional trial designs<sup>60</sup>. This has been viewed to create a challenging, yet fast-paced work environment with many career development opportunities<sup>60</sup>. | there is no end point<sup>60</sup>. Uncertain staffing costs<sup>43</sup> as adaptations from accumulating data may change recruitment needs or treatment arms that staff need to be trained to deliver, or trial amendments can lead to issues with ensuring adequate and timely access to intervention supplies<sup>24</sup>. Technical staff considerations include the considerable skills needed to complete trial simulations for different scenarios prior to starting a trial<sup>9</sup> as was required for trials such as AVERT-DOSE<sup>36</sup> and STEP<sup>39</sup>. Additional statistical staff may also be required for interim analyses, protocol development and statistical analysis planning and execution<sup>43</sup>. |
| **Ethical** |  |  |
| Ethical approval | Adaptive clinical trial designs have been viewed to pose few ethical disadvantages from a societal perspective<sup>56</sup>. | Communicating design aspects that may change down the track via study documents such as the Participant Information Sheet can pose challenges<sup>24</sup>. Concerns have also been raised about whether ethical review boards are ready to accept adaptive designs<sup>57</sup>, although the many completed and ongoing adaptive trials may counterbalance this concern. Manifold governance questions when dealing with multiple medicinal products or medical devices can discourage researchers working in small and moderate sized Universities<sup>61</sup>. |
| Changes mid trial, including stopping early based on reviewing data | Allows informative and more ethical decisions to be made in a timely manner during the trial e.g., increasing a participant's probability of being randomised into an arm that is higher performing<sup>62</sup>. Reduces the number of patients exposed to unnecessary risk of an ineffective treatment and allow them the opportunity to explore more promising alternatives<sup>5, 62</sup>. | An adaptive change to a trial design may lead to results after the adaptation that are different from those before the adaptation<sup>5</sup>. Knowledge of comparative interim results by trial management personnel in cases of unblinding, may make it difficult for regulators to determine whether, for example, a protocol amendment seemingly well-motivated by information external to the trial was influenced, in any way, by access to accumulating comparative data. Also, changing rules in an ongoing trial can also create complications with regulators.<sup>63</sup> |
| **Scientific** |  |  |
| Statistical efficiency | May increase the chance to detect a true effect (greater statistical power) than non-adaptive trials, or may provide the same statistical power with a smaller expected sample size or shorter expected trial | The opportunity for efficiency gains may be limited by important scientific constraints or in certain clinical settings<sup>5</sup>, e.g., when a minimum sample size is expected for a reliable evaluation of other outcomes (safety or secondary endpoints) or when the primary |
|  | duration<sup>5</sup>. | outcome of interest is ascertained over a longer window than it takes to enroll most or all patients in the trial (prevention or delayed outcome)<sup>5</sup>. |
| Design complexity | Access to statistical support ensures appropriate analytical methods are used to preserve trial validity and integrity<sup>1</sup>. | Widely recognized as more complex than traditional fixed designs<sup>1, 2, 5, 24, 62, 64</sup>, which can increase trial costs<sup>43</sup>. |
| Understanding of treatment effects | Can answer broader questions generally not feasible with non-adaptive designs e.g., adaptive enrichment enables efficacy/effectiveness in targeted subgroups of a population to be demonstrated where non-adaptive designs would require infeasibly large sample sizes<sup>5</sup>. | Inadequate access to statistical design support can increase the chance of erroneous conclusions and introduce bias in estimates.<sup>5</sup> |
| **Regulatory** |  |  |
| Regulatory discussions | Not the intent of regulatory bodies e.g., US Food and Drug Administration (FDA)<sup>5</sup> or European Medicines Agency (EMA)<sup>65</sup> to require or restrict the use of adaptive designs in general or specific settings. | The increased complexity of some adaptive trials and uncertainties regarding their operating characteristics may warrant earlier and more extensive interactions with regulatory agencies than usual<sup>5</sup>. |
| Regulatory evaluations | Additional opportunities for review may help to maintain (or even enhance) trial integrity and validity<sup>5</sup>. | Review of complex adaptive designs often involves challenging evaluations of design operating characteristics, usually requiring extensive computer simulations as well as increased discussion across disciplines and regulatory offices about the evaluations<sup>5</sup>. Regulatory agencies tend to review proposals for adaptive designs with greater scrutiny than they give to conventional designs<sup>62</sup>. This arises from serious concern poorly conceived designs may not control the type I error and may actually be less efficient than conventional designs<sup>62</sup>. |

### Clinical, research and technical workforce readiness

Adaptive clinical trials may impact research and clinical workforce readiness. Adaptive trials require access to statistical, data and trial management staff who are fluent in their design^42^ to manage the potential impact on trial costs. A mock-costing exercise completed with seven UK Clinical Trial Units estimated non-platform adaptive trials with pharmacologic interventions were 2-4% more expensive than a non-adaptive trial equivalent^43^. The highest increase was for statistical staff, with lower increases for data and trial management staff^43^. Costings for appropriate statistical staff are highly dependent on the nature of the adaptive feature(s) included^42^. During trial conduct, adaptations that occur at prespecified interim analyses (e.g., sample size reestimation, multi-arm, multi-stage) may require more intensive ongoing input than adaptations that are managed on a day-to-day basis by software that implement methods to adaptively randomise individual participants (e.g., covariate-adjusted, response-adaptive). However, such randomisation methods often require bespoke software^36, 39, 44^ to be developed prior to trial start for which the code is generally not publicly available^45^. It is important to note such development skills are different to those required to perform trial design and analysis work reflecting a unique workforce requirement for some adaptive designs.

Adaptive designs can impact site clinical and trial management staff. There can be additional costs associated with supplementary training in adaptive trial processes and procedures, and time to gain consent may be longer due to the complexity of transparently discussing adaptive features with potential participants^42^. Countering potential increased costs, trials that can be stopped for futility can create efficiencies that traditional trials cannot offer. For example, an adaptive platform trial allows the same participant to be enrolled in multiple distinct intervention trials, thereby answering many questions through participation in only one trial^46^. This may mitigate challenges for patients in choosing between trials and clinicians maintaining recruitment across competing trials. Some adaptive features (e.g., adaptive sample size reestimation, changes in intervention arms open to randomisation^47^) can lead to uncertainty in the amount of site staff required and result in short-term contracts to ensure adaptive features (whether enacted or not) can be accommodated. Other features that are more enduring (e.g., adaptive platform) may offer greater stability for trial staff due to their perpetual nature^46^. As such, the impact on the site workforce varies depending on the adaptive features included in a trial, but may also be impacted by the volume of trials operating at a site, whether the site is part of a network (e.g., NIH StrokeNet) or has an accessible Clinical Trial Unit (e.g., UK centres), and experience of the site^24, 42^. These issues have been largely acknowledged in reports from high-income countries and may be more pronounced in low-middle income countries and rural/regional centres.

With growing uptake of adaptive features (Table 1), it is important to consider the role of professional organisations (e.g., World Stroke Organisation, European Stroke Organisation) and networks (e.g., CanStroke Recovery Trials, Canadian Stroke Consortium, Australasian Stroke Trials Network, NIH StrokeNet^48^, Global Alliance of Independent Networks focused on Stroke trials (GAINS), Indian Stroke Clinical Trial Network (INSTRuCT^49^)), European Stroke Organsiation Trial Alliance (ESOTA^50^)) to prepare current and future generations of clinical trialists to embrace, where appropriate, adaptive trial designs. Clinical trial stakeholders have identified that broadening the acceptance and uptake of adaptive trials will depend on access to training, guidelines, and toolkits to ensure proper use of adaptive trial designs in practice^51^. Currently, most educational opportunities (e.g., webinars, scientific meetings) do not provide the depth of training required. This highlights a critical training gap that needs to be addressed in the immediate future.

## Conclusions

Our scoping review underpins consolidated knowledge about the use of adaptive trials by examining stroke as the exemplar neurological condition. There is considerable use of adaptive design features across efficacy and effectiveness trials of acute stroke treatments, and emerging use for trials of rehabilitation or prevention. The trial examples highlighted how meaningful decisions concerning the use of adaptive features can be applied in stroke to deliver more efficient and ethical trials, which may in turn deliver cost-savings. From a public health perspective, adaptive trial designs can no longer be overlooked and present an efficient and ethical trial design approach for national funding agencies to support. When adaptive features are incorporated, it is imperative that they are used correctly to preserve trial validity and integrity. Inclusion of any adaptive feature(s) in a clinical trial is no panacea. Researchers, funders, and sponsors will always need to carefully consider if an adaptive design provides the most appropriate approach to answer the research question, just as with any suite of design tools. Time and resources must be allocated to conduct the preparation work required to design an adaptive trial. Once doing an adaptive trial, comprehensive and transparent reporting of all adaptive features across all trial sources (i.e., trial publications and registration) is essential. There is scope to improve reporting across sources based on the number of trials identified through authorship knowledge of the field rather than through explicit reporting in publications and registries. Adherence to available guidelines (i.e., CONSORT ACE^1^) will enhance quality of reporting and ability for others to find and learn from prior adaptive trials. In line with this, greater knowledge across the research workforce will support peer-review of grants and publications that include adaptive design features. Overall, we have demonstrated that adaptive trials can be designed, funded, conducted, and published for a wide range of stroke research questions. We advocate for the broader, responsible adoption of adaptive designs to address many amenable clinical research questions in a more efficient and flexible manner.

## Data Availability

All data produced in the present work are contained in the manuscript

## Acknowledgements

Nil.

