## Supplemental Materials for "Adaptive trials in stroke: Current use & future directions"

| | |
|---|---|
| **Title** | Adaptive trials in stroke: Current use & future directions |
| **Short title** | Adaptive trial methods in stroke |


**<u>Corresponding author:</u>**

Dr Kathryn Hayward

Level 7 Alan Gilbert Building

University of Melbourne

161 Barry Street

Carlton Victoria Australia 3050


**Contents:**


A) Supplemental Methods

B) Supplemental Results

C) Supplemental Table 1

D) Supplemental Table 2

E) Supplemental References

### A. Supplemental Methods for scoping review of literature

Focusing on the recent use of adaptive features in stroke trials, we scoped over a five-year period (1 January 2018 to 2 May 2023) for ongoing and completed adaptive clinical trials via trial registries (Australian and New Zealand Clinical Trial Registry and clinicaltrials.gov) and publications (protocols and completed trials) via PubMed Central. Search terms of stroke and trial* and (adapt* or Bayesian or platform), as well as adaptive feature specific words from the CONSORT ACE statement[1] were used. The search strategy was broad as underreporting of adaptive features in clinical research across medicine has been documented[3, 4]. All results were independently reviewed by two authors (KH/ED) using Excel (registry results) or Covidence (PubMed results), and conflicts resolved through discussion with a third reviewer (LC). Trial investigators were contacted in cases of conflicting evidence of eligibility (e.g., mismatch between registration and protocol). Eligible clinical trials (ongoing [registered trial or published protocol] or completed) enrolled people with stroke to receive an acute, rehabilitation or preventative stroke treatment via a clinical trial that included at least one adaptive feature (Table 1) to answer an efficacy or effectiveness research question (i.e., phase II, phase III, seamless phase I-II or IIb-III). A particular feature was deemed present if it existed on its own rather than as a biproduct of another feature e.g., group sequential may result in a change in sample size, but such a change is not the purpose for using this feature. Trial information was linked where able i.e., registration, protocol, and results papers, and status updated if trial results became available. Preclinical stroke trials were excluded. Identified reviews and studies that simulated adaptive designs or reanalysed a trial using an adaptive design approach were used to inform a discussion about the utility of adaptive designs. Reference lists of included studies were hand searched and authorship knowledge of the field was used to identify additional stroke trials (published or registered trials) and evidence to inform a discussion of adaptive trials in stroke and neurology more generally. Data was independently extracted by one author (ED) and verified by another author (KH/BCVC). Conflicts were resolved through discussion with an additional independent author (KH/BCVC/LC). Data were collated and descriptively summarised.

### B. Supplemental Results of scoping review

From 445 trial registry and 1291 PubMed records, 29 trials (13 via trial registry, 16 via PubMed) were identified that enrolled people with stroke and included at least one adaptive design feature[4-27]. Sixteen additional trials were identified from reference lists [6], and authorship knowledge of the field (published[12, 14, 28-33]; registered: NCT04454788, NCT05047172, NCT05484154, NCT04167527, NCT03936361, NCT03907046, ACTRN12618001432202). Such trials were not identified through the primary searches as they did not reference adaptive features in the title/abstract/keywords.

C. **Supplemental Table 1: Completed adaptive clinical trials in stroke, n=14.**

| Short title(ref)<br>Year<br>Country/ies<br>Income status[a]<br>Support[b] | Source(s) | Population[c]<br>Treatment type[d]<br>Enrollment[e]<br>Primary endpoint<br>Registration[f] | Design | Adaptive feature(s) specified at design stage[g] | Details of if adaptive feature(s) were **enacted** or not enacted during the trial. |
|---|---|---|---|---|---|
| DAWN[4, 5]<br>2018<br>USA, Australia, Canada, France, Spain<br>High income<br>Industry-sponsored | ⊗ Registration<br>⊗ Protocol<br>⊗ Results | Acute<br>Device<br>Hyperacute<br>90d post<br>NCT02142283 | Multicentre, prospective, randomised, open-label trial with a Bayesian adaptive–enrichment design and blinded end point assessment. | ⊗ **Group sequential**<br>○ Sample size reestimation<br>⊗ **Patient population**<br>○ Treatment arm selection<br>○ Patient allocation: CAR<br>○ Patient allocation: RAR<br>○ Endpoint selection<br>○ Multi-arm, Multi-stage | **Group sequential:** Trial stopped due to results of a prespecified interim analysis 31m into enrollment. This was the first prespecified interim analysis that permitted stopping for this reason and was based on enrollment of 200 patients.<br>Patient population: Enrichment had not been triggered, which meant the analysis included the full population, regardless of infarct volume. |
| DEFUSE-3[6, 7]<br>2018<br>USA<br>High income<br>Investigator-initiated | ⊗ Registration<br>⊗ Protocol<br>⊗ Results | Acute<br>Device<br>Hyperacute<br>90d post<br>NCT02586415 | Randomised, open-label trial with blinded outcome assessment. Trial planned to use an adaptive enrichment design. | ⊗ **Group sequential**<br>○ Sample size reestimation<br>⊗ **Patient population**<br>○ Treatment arm selection<br>○ Patient allocation: CAR<br>○ Patient allocation: RAR<br>○ Endpoint selection<br>○ Multi-arm, Multi-stage | **Group sequential:** Enrollment was placed on hold, and an early interim analysis, including subgroup analysis of the primary and secondary efficacy outcomes in patients who would have been eligible for the DAWN trial, was requested by the sponsor. As a result of that interim analysis, the trial was halted because the prespecified efficacy boundary (P<0.0025) had been exceeded.<br>Patient population: No enrichment occurred. |
| EXTEND-IA TNK[8, 9]<br>2018 | ⊗ Registration<br>⊗ Protocol<br>⊗ Results | Acute<br>Pharmacological<br>Hyperacute | Multicentre, prospective, randomised, | ○ Group sequential<br>⊗ **Sample size reestimation**<br>○ Patient population | **Sample size reestimation:** Estimated sample size minimum of 120 and maximum of 276 patients. Prespecified to occur at |

| | | | | | |
|---|---|---|---|---|---|
| Australia, NZ<br>High income<br>Investigator-initiated, industry-supported | | 24h post<br>NCT02388061 | open-label, blinded-outcome trial. | ○ Treatment arm selection<br>○ Patient allocation: CAR<br>○ Patient allocation: RAR<br>○ Endpoint selection<br>○ Multi-arm, Multi-stage | 100 patients. Reestimation determined final sample size of 202 patients. |
| EXTEND[10, 11]<br>2019<br>Australia, Finland, NZ, Taiwan<br>High income<br>Investigator-initiated | ⊗ Registration<br>⊗ Protocol<br>⊗ Results | Acute<br>Pharmacological<br>Hyperacute<br>90d post<br>ACTRN12610000011088, NCT00887328/01580839 | Multicentre, randomised, placebo-controlled trial. | ○ Group sequential<br>⊗ **Sample size reestimation**<br>○ Patient population<br>○ Treatment arm selection<br>○ Patient allocation: CAR<br>○ Patient allocation: RAR<br>○ Endpoint selection<br>○ Multi-arm, Multi-stage | **Sample size reestimation:** Estimated sample size of 400. Reestimation performed from the first 200 patients. Analysis confirmed a final intended sample size of 310 patients. After 225 of the planned 310 patients had been enrolled, the trial was terminated because of a loss of equipoise after the publication of positive results from a previous trial (WAKE-UP). |
| SHINE[12, 13]<br>2019<br>USA<br>High income<br>Investigator-initiated | ⊗ Registration<br>⊗ Protocol<br>⊗ Results | Acute<br>Pharmacological<br>Hyperacute<br>90d post | RCT with blinded outcome assessment. | ⊗ **Group sequential**<br>⊗ **Sample size reestimation**<br>○ Patient population<br>○ Treatment arm selection<br>○ Patient allocation: CAR<br>⊗ **Patient allocation: RAR**<br>○ Endpoint selection<br>○ Multi-arm, Multi-stage | **Group sequential:** The first interim analysis was planned to be conducted once 500 patients reached the 90-day primary outcome. Subsequent interim analyses were to occur after 700, 900, and 1100 patients reached the 90-day primary outcome using the O'Brien and Fleming-type stopping guidelines. Stopping the trial due to harm would be considered if at any point the 2-sided 95% CI for the unadjusted relative risk for death excluded 1, or if the unadjusted absolute risk difference for the proportion of patients with severe hypoglycemia exceeded 4%. Study enrollment was stopped after the fourth prespecified interim analysis met the futility criteria.<br>**Sample size reestimation:** Blinded sample size reestimation was conducted prior to the first interim analysis. The plan was |

| | | | | | |
|---|---|---|---|---|---|
| | | | | | in place to avoid an underpowered trial if the assumed control proportion was incorrect.<br>**RAR:** Balanced based on baseline stroke severity, initiated or planned use of intravenous tissue plasminogen activator therapy, and clinical site. |
| TELEREHAB-2[14]<br>2019<br>USA<br>High income<br>Investigator-initiated | ⊗ Registration<br>○ Protocol<br>⊗ Results | Rehabilitation<br>Behavioural<br>Early subacute to chronic<br>30d post treatment<br>NCT02360488 | Randomized, assessor-blinded, noninferiority trial. | ○ Group sequential<br>○ Sample size reestimation<br>○ Patient population<br>○ Treatment arm selection<br>⊗ **Patient allocation: CAR**<br>○ Patient allocation: RAR<br>○ Endpoint selection<br>○ Multi-arm, Multi-stage | **CAR:** Used to ensure that values for time post-stroke, severity of impairment, age, enrollment site, and stroke subtype were balanced across the two treatment groups. |
| SOS[15, 16]<br>2020<br>USA<br>High income<br>Investigator-initiated | ⊗ Registration<br>⊗ Protocol<br>⊗ Results | Prevention<br>Behavioural<br>Not defined<br>6m post intervention<br>NCT02531074 | Pilot, prospective, randomised controlled trial with open blinded end point study. | ○ Group sequential<br>○ Sample size reestimation<br>○ Patient population<br>○ Treatment arm selection<br>⊗ **Patient allocation: CAR**<br>○ Patient allocation: RAR<br>○ Endpoint selection<br>○ Multi-arm, Multi-stage | **CAR:** Participants randomly assigned in a 1:1 ratio using adaptive covariate randomisation algorithm which used the Pearson chi-square statistic to measure treatment imbalances in stroke severity (mRS score of 0-1 or 2-3), age, and sex. |
| TST[17, 18]<br>2020<br>France, South Korea<br>High income | ⊗ Registration<br>⊗ Protocol<br>⊗ Results | Prevention<br>Pharmacological<br>Early subacute<br>Composite endpoint 6m<br>NCT01252875 | Randomised, parallel-group, event-driven trial. | ⊗ **Group sequential**<br>○ Sample size reestimation<br>○ Patient population<br>○ Treatment arm selection<br>○ Patient allocation: CAR<br>○ Patient allocation: RAR | **Group sequential:** No interim analysis initially planned since the two strategies considered as usual care. The steering committee decided to amend the plan and introduced an intermediate analysis when 65% of the primary endpoints were reached. For futility, the trial steering committee fixed a conditional power threshold value of 30%, and for efficacy |

| | | | | | |
|---|---|---|---|---|---|
| Investigator-initiated, industry-supported | | | | ○ Endpoint selection<br>○ Multi-arm, Multi-stage | (superiority) a nominal value of 0·0076 (O'Brien Fleming) for the intermediate analysis and equal to 0·0476 for the final analysis. |
| CPASS[19, 20]<br>2021<br>USA<br>High income<br>Investigator-initiated | ⊗ Registration<br>⊗ Protocol<br>⊗ Results | Rehabilitation<br>Behavioural<br>Early subacute<br>1y after stroke<br>NCT02235974 | Randomised controlled trial. | ○ Group sequential<br>○ Sample size reestimation<br>○ Patient population<br>○ Treatment arm selection<br>⊗ **Patient allocation: CAR**<br>○ Patient allocation: RAR<br>○ Endpoint selection<br>○ Multi-arm, Multi-stage | **CAR:** Participants adaptively randomised into one of four study groups by the study statisticians after informed consent and baseline assessment data were entered into a database. Groups balanced with respect to participant age, number of days from stroke onset to baseline, stroke type, baseline ARAT score, arm concordance, and overall stroke severity. |
| Qizhitongluo[21]<br>2021<br>China<br>Upper Middle income<br>Investigator-initiated | ⊗ Registration<br>○ Protocol<br>⊗ Results | Rehabilitation<br>Pharmacological<br>Early subacute<br>Post-intervention<br>NCT01762163 | Randomised, multicentre, double-blind, placebo- and active-controlled trial. | ⊗ **Group sequential**<br>○ Sample size reestimation<br>○ Patient population<br>○ Treatment arm selection<br>○ Patient allocation: CAR<br>○ Patient allocation: RAR<br>○ Endpoint selection<br>○ Multi-arm, Multi-stage | **Group sequential:** A total of 624 participants needed to be enrolled. An interim analysis with the use of O'Brien-Fleming stopping boundaries was performed to re-estimate the sample size after 312 patients were recruited. Based on the interim analysis and advice of the data management committee, the trial continued, and the sample size remained the initial size. |
| SWIFT-DIRECT[22, 23]<br>2022<br>Austria, Finland, France, Germany, Spain, Switzerland, UK & Canada | ⊗ Registration<br>⊗ Protocol<br>⊗ Results | Acute<br>Device<br>Hyperacute<br>90d after randomisation<br>NCT03192332 | A multicentre, prospective, randomised, open-label, blinded-endpoint trial utilizing an adaptive statistical design. | ⊗ **Group sequential**<br>⊗ **Sample size reestimation**<br>○ Patient population<br>○ Treatment arm selection<br>○ Patient allocation: CAR<br>○ Patient allocation: RAR<br>○ Endpoint selection<br>○ Multi-arm, Multi-stage | **Group sequential:** After 202 patients reached the primary outcome at 90d, an interim analysis was conducted, including (re)assessments of sample size and conditional power. Stopping for futility based on conditional power was conservative concerning the type1-error rate.<br>**Sample size reestimation:** Sample size was recalculated using the observed functional independence in the combined group as reference. The targeted sample size could be |

| | | | | | |
|---|---|---|---|---|---|
| High income<br>Investigator-initiated, industry-supported | | | | | increased if the recalculated sample size was larger than the initial one (i.e., if the observed functional independence was closer to 50% than the assumed 62%). The re-estimated sample size was lower than the initial one and no adjustment was made. |
| SELECT-2[24, 25]<br>2023<br>USA, Canada, Europe, Australia, NZ<br>High income<br>Investigator-initiated, industry-supported | ⊗ Registration<br>⊗ Protocol<br>⊗ Results | Acute<br>Device<br>Hyperacute<br>90d post onset<br>NCT03876457 | Randomised, open-label clinical trial with adaptive enrichment design and blinded end-point assessment. | ⊗ **Group sequential**<br>○ Sample size reestimation<br>⊗ **Patient population**<br>○ Treatment arm selection<br>○ Patient allocation: CAR<br>○ Patient allocation: RAR<br>○ Endpoint selection<br>○ Multi-arm, Multi-stage | **Group sequential:** Original trial design included two prespecified interim analyses, which were to be performed when 200 patients and 380 patients enrolled in the trial had completed the 90d follow-up. The DSMB reviewed the results of the first interim analysis of the data for 200 patients and recommended that the trial continue. In light of the RESCUE-Japan LIMIT trial, the board requested to review the data for the efficacy and safety outcomes after 300 patients enrolled in the trial had completed 90d follow-up. This review showed that the prespecified efficacy boundary had been crossed in favour of EVT. Board recommended that enrollment be stopped.<br>Patient population: No enrichment occurred. |
| SPRINT India[26, 27]<br>2023<br>India<br>Lower middle income<br>Investigator-initiated | ⊗ Registration<br>⊗ Protocol<br>⊗ Results | Prevention<br>Behavioural<br>Acute-early subacute<br>Composite endpoint at 1y<br>NCT03228979 | Multicentre, randomised, parallel-design, adaptive and blinded end-point clinical trial of sub-acute stroke patients. | ⊗ **Group sequential**<br>○ Sample size reestimation<br>○ Patient population<br>○ Treatment arm selection<br>○ Patient allocation: CAR<br>○ Patient allocation: RAR<br>○ Endpoint selection<br>○ Multi-arm, Multi-stage | **Group sequential:** An interim futility analysis was planned after 50% enrolment or occurrence of 50% of events, whichever came first. However, owing to the pandemic, the interim analysis was done at 67% of recruitment. The difference between groups in the proportion of patients with events at 1y was taken as the endpoint to calculate the conditional power. The trial was stopped for futility by the DSMB because of fewer vascular events than expected and low conditional power. |

| | | | | | |
|---|---|---|---|---|---|
| TENSION[34, 35]<br>2023<br>Austria, Czech Republic, Denmark, France, Germany, Norway, Slovakia, Spain, Canada<br>High income<br>Investigator-initiated | ⊗ Registration<br>⊗ Protocol<br>⊗ Results | Acute<br>Device<br>Hyperacute<br>90d post onset<br>NCT03094715 | Randomised controlled, open label, blinded endpoint, two-arm, post-market study with interim analyses (pre-specified stopping rules). | ⊗ **Group sequential**<br>O Sample size reestimation<br>O Patient population<br>O Treatment arm selection<br>O Patient allocation: CAR<br>O Patient allocation: RAR<br>O Endpoint selection<br>O Multi-arm, Multi-stage | The study included two interim analyses after a third and two-thirds of the patients had completed the 90-day follow-up, for futility and early efficacy. The trial was stopped early for efficacy after the pre-planned first interim analysis of 222 patients. The primary outcome analysis showed a significant shift towards an improved functional outcome across the entire range of the mRS at 90 days in patients assigned to endovascular thrombectomy. |

Notes: [a]Income status from The World Bank https://datatopics.worldbank.org/world-development-indicators/the-world-by-income-and-region.html defined as either high-income, upper-middle income, lower-middle income, low-income. [b]Support defined via trial sponsorship and funding definitions across sources as industry-sponsored, investigator-initiated industry-supported, investigator-initiated. Industry supported means that there was substantive involvement such as supply of drug or device, while the funding may have originated from a government body (such as NIH, NHMRC). [c]Population defined as acute, rehabilitation, or prevention. [d]Treatment type defined as pharmacological, device or behavioural. [e]Timepoint of enrolment timepoints defined using the Stroke Recovery and Rehabilitation Roundtable recommendations[36] of 0-24h hyperacute, 25h-7d acute, >7-90d early subacute, >90-180d late subacute and beyond 180d chronic. [f]Registration NCT=clinicaltrials.gov, ACTRN=Australian and New Zealand Clinical Trial Registry. [g]Adaptations to patient allocation CAR = covariate-adaptive randomisation. Adaptations to patient allocation RAR = response-adaptive randomisation. AIS=acute ischaemic stroke. DSMB=data and safety monitoring board. ET=endovascular thrombectomy. ICH=intracerebral haemorrhage. LVO=large vessel occlusion. mRS=modified Rankin Scale. TIA=transient ischaemic attack. MT=mechanical thrombectomy. y=years. m=months. w=weeks. d=days. h=hours. min=minutes.

D. **Supplemental Table 2: Ongoing adaptive clinical trials in stroke with a published protocol, n=15 and trial registration only, n=16.**

| Short title(ref)<br>Year (published/registered)<br>Country/ies<br>Income status[a]<br>Support[b] | Source(s) | Population[c]<br>Treatment type[d]<br>Enrollment[e]<br>Primary endpoint<br>Registration[f] | Design | Adaptive feature(s)[g] | Details of conditions that determine how the adaptive feature(s) are enacted |
|---|---|---|---|---|---|
| Adaptive trials with a published protocol, n=15 | | | | | |
| TEXAIS[37]*<br>2018<br>Australia<br>High income<br>Investigator-initiated | ⊗ Registration<br>⊗ Protocol<br>○ Results | Acute<br>Pharmacological<br>Hyperacute<br>7d post onset<br>ACTRN12617000409370 | Multicentre, prospective, randomised, open label, blinded end-point trial with pre-planned adaptive sample size re-estimation. | ○ Group sequential<br>⊗ **Sample size reestimation**<br>○ Patient population<br>○ Treatment arm selection<br>○ Patient allocation: CAR<br>○ Patient allocation: RAR<br>○ Endpoint selection<br>○ Multi-arm, Multi-stage | Conditional power will be evaluated at an interim analysis, and if within the pre-specified promising zone, the sample size should be increased, subject to a pre-determined upper limit (650 patients). |
| ARCADIA[33]<br>2019<br>USA, Canada<br>High income<br>Investigator-initiated, industry-supported | ⊗ Registration<br>⊗ Protocol<br>○ Results | Prevention<br>Pharmacological<br>Hyperacute, acute, early subacute, late subacute and chronic<br>Recurrent stroke<br>NCT03192215 | Multicenter, biomarker-driven, randomized, double-blind, active-control, phase 3 clinical trial | ⊗ **Group sequential**<br>○ Sample size reestimation<br>○ Patient population<br>○ Treatment arm selection<br>○ Patient allocation: CAR<br>○ Patient allocation: RAR<br>○ Endpoint selection<br>○ Multi-arm, Multi-stage | One interim analysis for efficacy (or harm) and futility anticipated at the halfway point of the trial. An O'Brien-Fleming type Lan-DeMets error spending function will be used for the interim analysis boundaries, assuming a nonbinding futility boundary. |
| START[38]<br>2019<br>USA | ⊗ Registration<br>⊗ Protocol<br>○ Results | Acute<br>Pharmacological<br>Acute | Prospective, multicentre, randomised, | ⊗ **Group sequential**<br>○ Sample size reestimation<br>○ Patient population | RAR will be used allocate subjects to the four time-to-treatment arms independently for each level of risk. The first 100 subjects will be equally |

| | | | | | |
|---|---|---|---|---|---|
| High income<br>Investigator initiated | | 30d of index stroke<br>NCT03021928 | response-adaptive, multi-arm time-to-treatment trial. | ○ Treatment arm selection<br>○ Patient allocation: CAR<br>⊗ **Patient allocation: RAR**<br>○ Endpoint selection<br>○ Multi-arm, Multi-stage | randomised in a 1:1:1:1 ratio to each cohort. After initial 100 enrollments, allocation ratios will be adjusted through interim analyses every 100 enrollments. |
| Sleep-SMART[32]<br>2020<br>USA<br>High income<br>Investigator-initiated | ⊗ Registration<br>⊗ Protocol<br>○ Results | Prevention and recovery<br>Pharmacological<br>Acute & early subacute<br>Composite endpoint 6m post stroke (prevention) and 3m post stroke (recovery)<br>NCT03812653 | Phase III, multicentre, prospective randomized, open, blinded outcome event assessed controlled trial. | ⊗ **Group sequential**<br>⊗ **Sample size reestimation**<br>○ Patient population<br>○ Treatment arm selection<br>○ Patient allocation: CAR<br>⊗ **Patient allocation: RAR**<br>○ Endpoint selection<br>○ Multi-arm, Multi-stage | After 40% of subjects have adjudicated prevention outcomes available, one interim assessment of the prevention outcome for both efficacy and futility, and one assessment of the recovery outcome only for overwhelming efficacy will be performed. If stopping rules are met for either outcome, the DSMB will discuss potential protocol modifications to allow completion of the other aim. Plan to conduct a blinded sample size re-estimation at the time of the single interim analysis. Randomization (1:1) will be performed centrally via a web-based system, using a minimal sufficient balance approach that prevents serious imbalances in the following important baseline covariates: site, age, NIHSS, and use of intravenous thrombolysis or attempted endovascular therapy. |
| MOST[39]*<br>2021<br>USA<br>High income<br>Investigator-initiated | ⊗ Registration<br>⊗ Protocol<br>○ Results | Acute<br>Pharmacological<br>Hyperacute<br>90d post randomisation<br>NCT03735979 | Three-armed, adaptive, single blind randomised controlled clinical trial conducted. | ⊗ **Group sequential**<br>○ Sample size reestimation<br>○ Patient population<br>○ Treatment arm selection<br>○ Patient allocation: CAR<br>⊗ **Patient allocation: RAR** | Interim analyses are planned to occur after 500, 700, and 900 subjects have been randomised. The predictive probability of a successful final analysis has been calculated based on different assumptions about the remaining subjects to be enrolled. When n=500, one (or both) arm(s) may be stopped for |

| | | | | | |
|---|---|---|---|---|---|
| | | | | ○ Endpoint selection<br>○ Multi-arm, Multi-stage | futility if there is less than 20% probability of demonstrating benefit in either intervention if the trial were to continue. Next, one (or both) arm(s) may be stopped for futility when n=700 or n=900 if there is less than 5% probability of demonstrating benefit in either intervention if the trial were to continue. One (or both) arm(s) may be stopped early for efficacy after 700 or 900 subjects if an arm has an expected successful predictive probability of demonstrating superiority to control of at least 99%. |
| Restart TICrH[40]<br>2021<br>USA<br>High income<br>Investigator-initiated | ⊗ Registration<br>⊗ Protocol<br>○ Results | Acute<br>Pharmacological<br>Acute & early subacute<br>Composite endpoint 60d post stroke<br>NCT04891861 | Prospective randomised open label blinded endpoint response adaptive clinical trial. | ⊗ **Group sequential**<br>○ Sample size reestimation<br>○ Patient population<br>○ Treatment arm selection<br>○ Patient allocation: CAR<br>⊗ **Patient allocation: RAR**<br>○ Endpoint selection<br>○ Multi-arm, Multi-stage | Using a Bayesian response adaptive randomisation design with potential to stop early for success or futility, plan to enroll a maximum of 1100 subjects (expected 10% lost to follow-up, leaving 990 analysable). |
| DUMAS[41]<br>2022<br>Netherlands<br>High income<br>Investigator-initiated, industry-supported | ⊗ Registration<br>⊗ Protocol<br>○ Results | Acute<br>Pharmacological<br>Hyperacute<br>24h after study drug administered<br>NCT04256473 | Multicentre trial with a prospective randomised open-label blinded end-point design, and an adaptive design for dose optimization. | ⊗ **Group sequential**<br>○ Sample size reestimation<br>○ Patient population<br>○ Treatment arm selection<br>○ Patient allocation: CAR<br>○ Patient allocation: RAR<br>○ Endpoint selection<br>○ Multi-arm, Multi-stage | The first interim is planned to occur after 60 subjects with a discharge diagnosis of ischaemic stroke have data up to 48h. Subsequent interim analyses will be conducted after inclusion of every 20 patients with a discharge diagnosis of ischaemic stroke and will continue until full accrual. Since interims are defined by calendar time, the total number of planned interims is random and will depend on the rate at which subjects accrue to the |

| | | | | | |
|---|---|---|---|---|---|
| | | | | | trial. In the initial phase, mixed quantitative-qualitative review for safety will be carried out by the DSMB, after inclusion of every 10 patients. |
| FASTEST[29]<br>2022<br>US, Canada, Germany, Japan, Spain, UK<br>High income<br>Investigator-initiated, industry-sponsored | ⊗ Registration<br>⊗ Protocol<br>○ Results | Acute<br>Pharmacological<br>Hyperacute<br>180d<br>NCT03496883 | Randomised, placebo-controlled, double-blind, parallel group trial. | ⊗ **Group sequential**<br>○ Sample size reestimation<br>○ Patient population<br>○ Treatment arm selection<br>○ Patient allocation: CAR<br>○ Patient allocation: RAR<br>○ Endpoint selection<br>○ Multi-arm, Multi-stage | One interim analysis is planned after approximately one-half patients have been evaluated. An alpha-spending function with O'Brien-Fleming stopping boundaries (two-sided) will be used. |
| ReCAPS[42]<br>2022<br>Australia<br>High income<br>Investigator-initiated | ⊗ Registration<br>⊗ Protocol<br>○ Results | Prevention<br>Behavioural<br>Early subacute<br>90d post randomisation<br>ACTRN12618001468213 | Prospective, parallel two-group, double-blind, multisite, RCT with blinded outcome assessment and intention-to-treat analysis. | ○ Group sequential<br>⊗ **Sample size reestimation**<br>○ Patient population<br>○ Treatment arm selection<br>○ Patient allocation: CAR<br>○ Patient allocation: RAR<br>○ Endpoint selection<br>○ Multi-arm, Multi-stage | Estimated sample size of 890 participants (445 per group). Sample size will be adaptively reestimated when outcomes for 668 participants are obtained, with maximum sample capped at 1100. |
| STOP MSU[43]*<br>2022<br>Australia, Finland, New Zealand, Taiwan, Vietnam<br>High income, Lower middle income | ⊗ Registration<br>⊗ Protocol<br>○ Results | Acute<br>Pharmacological<br>Hyperacute<br>24h post randomisation<br>NCT03385928 | Double-blind, randomised, placebo-controlled, multicentre clinical trial, conducted within | ○ Group sequential<br>⊗ **Sample size reestimation**<br>○ Patient population<br>○ Treatment arm selection<br>○ Patient allocation: CAR<br>○ Patient allocation: RAR<br>○ Endpoint selection | Estimated sample size of 180 patients (90 per arm). Sample size will be adaptively reestimated based on the outcomes of 144 patients, with a possible increase to a prespecified maximum of 326 patients. |

| | | | | | |
|---|---|---|---|---|---|
| Investigator-initiated | | | the estimand statistical framework. | ○ Multi-arm, Multi-stage | |
| AVERT-DOSE[44]<br>2023<br>Australia, India, Malaysia, Singapore, UK, Brazil, Ireland<br>High income, Upper middle income, Lower middle income<br>Investigator-initiated | ⊗ Registration<br>⊗ Protocol<br>○ Results | Rehabilitation<br>Behavioural<br>Hyperacute-acute<br>3m post-stroke<br>ACTRN12619000557134 | Multi-arm, multi-stage, covariate adjusted, response adaptive, randomised trial. | ○ Group sequential<br>⊗ **Sample size reestimation**<br>○ Patient population<br>⊗ **Treatment arm selection**<br>⊗ **Patient allocation: CAR**<br>⊗ **Patient allocation: RAR**<br>○ Endpoint selection<br>⊗ **Multi-arm, Multi-stage** | In Stage 1 (mild or moderate severity strata), 25% of patients will be randomised into the reference arm, while randomisation into three intervention arms is guided by the adaptive algorithm. At Stage 2, randomisation into the reference arm and the single selected intervention arm will be guided by the adaptive algorithm. Covariates are stroke severity (stratified a priori), age, geographic care region, and reperfusion interventions, all used for adaptive adjustment. RAR algorithmically assigns new participants based on 3m outcomes of previous participants. Since all RAR procedures are dependent on estimation of unknown parameters, the 3m delay in response-adaptation reduces the possibility of major changes to recruitment patterns early in the trial and allows for a balanced number of participants to be recruited in all arms. |
| ENRICH[28]*<br>2023<br>USA<br>High income<br>Industry-sponsored | ⊗ Registration<br>⊗ Protocol<br>○ Results | Acute<br>Device<br>Hyperacute: within 24h after the onset of symptoms<br>180d post-randomisation<br>NCT02880878 | Adaptive, two-arm, randomised comparative effectiveness study. | ⊗ **Group sequential**<br>○ Sample size reestimation<br>⊗ **Patient population**<br>○ Treatment arm selection<br>○ Patient allocation: CAR<br>○ Patient allocation: RAR<br>○ Endpoint selection | Scheduled interim analyses, beginning after 150 patients have been enrolled, will be used to guide the adaptive sample size and study enrichment based on haemorrhage location. A maximum of 300 subjects will be enrolled. A trial update will be performed when the number of patients enrolled is equal to 150, 175, 200, 225, 250, and 275 patients. |

| | | | | | |
|---|---|---|---|---|---|
| | | | | ○ Multi-arm, Multi-stage | |
| TESLA[30]*<br>2023<br>USA, Europe (countries not defined)<br>High income<br>Investigator-initiated, industry-support | ⊗ Registration<br>⊗ Protocol<br>○ Results | Acute<br>Device<br>Hyperacute<br>3m post-randomisation<br>NCT03805308 | Prospective RCT, multicentre, open-label, assessor-blinded trial with adaptive enrichment design. | ○ Group sequential<br>○ Sample size reestimation<br>⊗ **Patient population**<br>○ Treatment arm selection<br>○ Patient allocation: CAR<br>○ Patient allocation: RAR<br>○ Endpoint selection<br>○ Multi-arm, Multi-stage | An adaptive enrichment design was incorporated for potential removal of ASPECTS 2 to 3 patients if they demonstrated futility. |
| TIMELESS[31]*<br>2023<br>Canada, USA<br>High income<br>Industry-sponsored | ⊗ Registration<br>⊗ Protocol<br>○ Results | Acute<br>Pharmacological<br>Hyperacute<br>90d post<br>NCT03785678 | Double-blind, randomised, placebo-controlled trial. | ⊗ **Group sequential**<br>○ Sample size reestimation<br>○ Patient population<br>○ Treatment arm selection<br>○ Patient allocation: CAR<br>○ Patient allocation: RAR<br>○ Endpoint selection<br>○ Multi-arm, Multi-stage | Two safety interim analyses are planned to occur when the first 25 and 50 patients have completed the 72-96h visit assessments. One efficacy interim analysis will be conducted when approximately 50% of the total patients have completed the 90d follow-up assessment. |
| TASTE[45]<br>2023<br>Australia, Taiwan, Vietnam<br>High income, Lower middle income<br>Investigator-initiated, industry-supported | ⊗ Registration<br>⊗ Protocol<br>○ Results | Acute<br>Pharmacological<br>Hyperacute: within 4.5h of symptom onset<br>3m post<br>ACTRN12613000243718 | Multicentre, prospective, randomised open-label blinded endpoint, controlled non-inferiority trial. | ○ Group sequential<br>⊗ **Sample size reestimation**<br>○ Patient population<br>○ Treatment arm selection<br>○ Patient allocation: CAR<br>○ Patient allocation: RAR<br>○ Endpoint selection<br>○ Multi-arm, Multi-stage | Planned to occur based on the results from the first 546 patients. The reestimation recommended an adaptive increase from the minimum of 728 patients to 832 patients as the final sample size for this study. The maximum sample size was maintained at a cap of 1024 patients. |
| Adaptive trials with registration only, n=16 | | | | | |

| | | | | | |
|---|---|---|---|---|---|
| MIDAS-2<br>2018<br>Australia<br>High income<br>Investigator-initiated | ⊗ Registration<br>○ Protocol<br>○ Results | Rehabilitation<br>Pharmacological<br>Late subacute-Chronic<br>56d post intervention<br>ACTRN12618000602224 | Multicentre, prospective, randomised, placebo-controlled, double-blind, parallel group, with an adaptive sample size reestimation. | ○ Group sequential<br>⊗ **Sample size reestimation**<br>○ Patient population<br>○ Treatment arm selection<br>○ Patient allocation: CAR<br>○ Patient allocation: RAR<br>○ Endpoint selection<br>○ Multi-arm, Multi-stage | Estimated sample size of 300 patients (150 per arm). Sample size will be adaptively reestimated based on the outcomes of 200 patients, with a possible increase to a prespecified maximum of 400 patients (Mehta and Pocock). |
| VESPUR<br>2018<br>Australia<br>High income<br>Investigator-initiated | ⊗ Registration<br>○ Protocol<br>○ Results | Rehabilitation<br>Behaviour<br>Acute-Early subacute<br>3m<br>ACTRN12618001432202 | Simon's 2-stage design. | ⊗ **Group sequential**<br>○ Sample size reestimation<br>○ Patient population<br>○ Treatment arm selection<br>○ Patient allocation: CAR<br>○ Patient allocation: RAR<br>○ Endpoint selection<br>○ Multi-arm, Multi-stage | Analysis has two stages:<br>1. Stage 1 functions as a screening stage to determine if there is some indication of effect, and if it is worthy to continue enrollment into either the early and/or late study<br>2. Both Stage 1 and Stage 2 determine if early or late intervention is worthy investigating in a subsequent comparative group trial. |
| ASPIRE<br>2019<br>USA<br>High income<br>Investigator-initiated | ⊗ Registration<br>○ Protocol<br>○ Results | Prevention<br>Pharmacological<br>Early-late subacute<br>Up to 3y<br>NCT03907046 | Randomised, double-blinded, clinical trial. | ⊗ **Group sequential**<br>○ Sample size reestimation<br>○ Patient population<br>○ Treatment arm selection<br>○ Patient allocation: CAR<br>○ Patient allocation: RAR<br>○ Endpoint selection<br>○ Multi-arm, Multi-stage | Interim analysis planned after two-thirds of primary outcome events. |

| | | | | | |
|---|---|---|---|---|---|
| ENDOLOW<br>2019<br>USA, Canada, Germany<br>High income<br>Investigator-initiated, industry-supported | ⊗ Registration<br>○ Protocol<br>○ Results | Acute<br>Device<br>Hyperacute<br>90d<br>NCT04167527 | Phase II/III, prospective, randomized, open-label, blinded-endpoint. | ⊗ **Group sequential**<br>⊗ **Sample size reestimation**<br>○ Patient population<br>○ Treatment arm selection<br>○ Patient allocation: CAR<br>○ Patient allocation: RAR<br>○ Endpoint selection<br>○ Multi-arm, Multi-stage | Adaptive two-stage design trial allowing for early stopping for efficacy or futility after the first stage or continuation with stage two including the option of recalculating the sample size. Interim analysis will be performed after the primary endpoint is available for a total of 175 randomized patients. |
| SATURN<br>2019<br>USA, Canada<br>High income<br>Investigator-initiated | ⊗ Registration<br>○ Protocol<br>○ Results | Prevention<br>Pharmacological<br>Acute<br>Up to 24m<br>NCT03936361 | Multicentre, pragmatic, prospective, randomized, open-label, and blinded end-point assessment clinical trial. | ⊗ **Group sequential**<br>○ Sample size reestimation<br>○ Patient population<br>○ Treatment arm selection<br>○ Patient allocation: CAR<br>○ Patient allocation: RAR<br>○ Endpoint selection<br>○ Multi-arm, Multi-stage | One interim analysis for overwhelming efficacy and futility, conducted according to O'Brien-Fleming stopping boundary, is planned; the number and timing of analysis can be altered according to DSMB request. |
| ETERNAL<br>2020<br>Australia<br>High income<br>Investigator-initiated | ⊗ Registration<br>○ Protocol<br>○ Results | Acute<br>Pharmacological<br>Hyperacute<br>90d<br>NCT04454788 | Prospective, randomised, open-label, blinded endpoint. | ○ Group sequential<br>⊗ **Sample size reestimation**<br>○ Patient population<br>○ Treatment arm selection<br>⊗ **Patient allocation: CAR**<br>○ Patient allocation: RAR<br>○ Endpoint selection<br>○ Multi-arm, Multi-stage | Patients will be randomized to treatment with either standard of care (no intravenous thrombolytic treatment or intravenous alteplase 0.9mg/kg) or intravenous tenecteplase (0.25mg/kg), using an adaptive covariate adjusted randomisation procedure to minimise imbalance on prespecified covariates. Estimated sample size of 740 patients (370 per arm). Sample size will be adaptively reestimated based on the outcomes of 592 patients, with a possible increase to a prespecified maximum of 1000 patients (Mehta and Pocock). |

| | | | | | |
|---|---|---|---|---|---|
| EVACUATE<br>2020<br>Australia<br>High income<br>Investigator-initiated | ⊗ Registration<br>○ Protocol<br>○ Results | Acute<br>Device<br>Hyperacute<br>6m<br>NCT04434807 | Prospective, randomised, open-label, blinded endpoint design with seamless phase IIb-III transition. | ○ Group sequential<br>⊗ **Sample size reestimation**<br>○ Patient population<br>○ Treatment arm selection<br>⊗ **Patient allocation: CAR**<br>○ Patient allocation: RAR<br>⊗ **Endpoint selection**<br>⊗ **Multi-arm, Multi-stage** | Seamless phase IIb to III transition is planned if the intermediate endpoint (successful haematoma evacuation) was met in analysis of the first 52 patients. Adaptive sample size re-estimation (Mehta and Pocock) will be performed when 160 patients have completed 6m follow-up (minimum sample size 240, maximum sample size 434). Randomisation uses the modified common scale minimum sufficient balance algorithm that improves the balance of key prognostic variables while maximizing the randomness of allocation. |
| ACTISAVE<br>2021<br>USA, Germany<br>High income<br>Industry-sponsored | ⊗ Registration<br>○ Protocol<br>○ Results | Acute<br>Pharmacological<br>Hyperacute<br>90d post<br>NCT05070260 | A randomised, double blind, multicentre, multinational, placebo controlled, parallel group, single dose, adaptive. | ⊗ **Group sequential**<br>⊗ **Sample size reestimation**<br>⊗ **Patient population**<br>○ Treatment arm selection<br>○ Patient allocation: CAR<br>○ Patient allocation: RAR<br>○ Endpoint selection<br>○ Multi-arm, Multi-stage | Adaptive seamless phase II/III study (respectively Part 1 and 2) with a possibility to stop after end of phase II interim analysis due to futility. **Adaptive selection of the population**: i.e., either continue to enroll in the overall population or restrict enrolment to the MT+ subgroup. **Adaptive increase in sample size** given the estimated difference between glenzocimab and placebo on each of the dual endpoints at the second interim analysis in the selected population. The statistical analysis plans will provide the details rules for claiming the trial futile, for selecting the population, and to consider an adaptive increase in the sample size, as well as the method to control the type I error rate. |
| CAPTIVA<br>2021 | ⊗ Registration<br>○ Protocol | Prevention<br>Pharmacological | Randomised, parallel | ○ Group sequential<br>○ Sample size reestimation | The first stage (safety analysis when first 450 subjects have completed 12m follow-up) will assess |

| | | | | | |
|---|---|---|---|---|---|
| USA<br>High income<br>Investigator-initiated, industry-supported | ○ Results | Acute-early subacute<br>1y<br>NCT05047172 | assignment, quadruple blinded. | ○ Patient population<br>○ Treatment arm selection<br>○ Patient allocation: CAR<br>○ Patient allocation: RAR<br>○ Endpoint selection<br>⊗ **Multi-arm, Multi-stage** | whether there is an excess of ICH or non-ICH major hemorrhage in the rivaroxaban or ticagrelor arms of the trial that could lead to an early termination of one or both of those arms. The second stage (futility analysis when 50% of primary outcome has been observed) will determine if the experimental arm(s) that progress to the second stage are superior to the clopidogrel arm for lowering the 1-year rate of the primary endpoint (ischemic stroke, ICH, or vascular death). |
| POST-ETERNAL<br>2021<br>Australia<br>High income<br>Investigator-initiated | ⊗ Registration<br>○ Protocol<br>○ Results | Acute<br>Pharmacological<br>Hyperacute<br>90d post<br>NCT05105633 | Multi-arm, multi-stage, prospective, randomised, open-label, blinded endpoint design with seamless phase IIb/III transition. | ⊗ **Group sequential**<br>⊗ **Sample size reestimation**<br>○ Patient population<br>○ Treatment arm selection<br>⊗ **Patient allocation: CAR**<br>○ Patient allocation: RAR<br>⊗ **Endpoint selection**<br>○ Multi-arm, Multi-stage | Seamless phase IIb/III trial with adaptive sample size recalculation. Phase IIb planned to use the surrogate outcome of recanalization without symptomatic ICH to establish whether proceeding to Phase II is warranted. If results from the first n= 202 patients meet success criteria, the trial will seamlessly convert to a phase 3 design using mRS 0-1 at 3m as the primary outcome. Minimum n=320 with interim sample size re-estimation at n=240, maximum sample n=688 using the Mehta and Pocock method. Covariate adjusted minimum sufficient balance randomisation will then be applied to control for age, NIHSS and time from onset-to-randomisation. |
| ReMEDY 2<br>2021<br>USA | ⊗ Registration<br>○ Protocol<br>○ Results | Acute<br>Pharmacological<br>Hyperacute | Randomised, double-blind, placebo-controlled | ⊗ **Group sequential**<br>⊗ **Sample size reestimation**<br>○ Patient population | An interim analysis will be conducted at the end of phase II after approximately 144 patients complete their 90d assessments. Additional patients will |

| | | | | | |
|---|---|---|---|---|---|
| High income<br>Industry-sponsored | | 90d post<br>NCT05065216 | phase II/III seamless adaptive, multi-centre study. | ○ Treatment arm selection<br>○ Patient allocation: CAR<br>○ Patient allocation: RAR<br>○ Endpoint selection<br>○ Multi-arm, Multi-stage | continue to be enrolled while the database and interim statistical analysis are prepared for review. A DSMB will review the interim data for efficacy and safety to determine if the trial proceeds to phase III. The adaptive feature of the study includes a formal interim assessment of efficacy and the possibility for increasing the sample size. The study is initially planned to enroll an additional 220 patients in phase III (N=364 total). If sample size re-estimation is deemed appropriate from the interim analysis, a maximum of 584 patients may be enrolled in phase III (N=728 total). This adaptive design is (inferentially) seamless because it continues from phase II to III without pausing enrollment and because all (N=364 to 728 total) patients will be included in the final statistical analysis. |
| EXTEND-IA DNASE<br>2022<br>Australia<br>High income<br>Investigator-initiated | ⊗ Registration<br>○ Protocol<br>○ Results | Acute<br>Pharmacological<br>Hyperacute<br>24h post-treatment<br>NCT05203224 | Bayesian Optimal phase II dose-finding umbrella trial. | ⊗ **Group sequential**<br>○ Sample size reestimation<br>○ Patient population<br>○ Treatment arm selection<br>○ Patient allocation: CAR<br>○ Patient allocation: RAR<br>○ Endpoint selection<br>○ Multi-arm, Multi-stage | Single arm versus objective performance criterion of 20% substantial reperfusion prior to ET based on the EXTEND-IA TNK trials. Patients will receive a single intravenous dose of dornase alfa (at either 0·125mg/kg, 0·25mg/kg, or 0·5mg/kg) in escalating tiers. |
| FIT4ME<br>2022 | ⊗ Registration<br>○ Protocol | Rehabilitation<br>Behavioural | Multicentre early-phase dose- | ⊗ **Group sequential**<br>○ Sample size reestimation | Once 20 participants (4 cohorts x 5 participants) are enrolled in a dose without escalation or de- |

| | | | | | |
|---|---|---|---|---|---|
| Australia<br>High income<br>Investigator-initiated | ○ Results | Acute-Subacute<br>8w post intervention<br>ACTRN12622001083785 (Mild)/12622001103752 (Moderate) | escalation study. A flexible Bayesian Optimal Interval phase I/II (BOIN12) design. | ○ Patient population<br>○ Treatment arm selection<br>○ Patient allocation: CAR<br>⊗ **Patient allocation: RAR**<br>○ Endpoint selection<br>○ Multi-arm, Multi-stage | escalation, the study arm is stopped, and that dose is deemed appropriate to test in a larger trial. Each study arm will have a minimum sample size of 20 and a maximum of 50. Design considers risks (safety/feasibility) and benefits (efficacy) when deciding to increase intervention dose and cohorts are subsequently allocated to a specific dose. |
| LATE-MT<br>2022<br>China<br>Upper middle income<br>Investigator-initiated | ⊗ Registration<br>○ Protocol<br>○ Results | Acute<br>Device<br>Acute<br>90d post<br>NCT05326932 | Multicentre, prospective, randomised, open-label, adaptive group sequential design, blinded endpoint assessment clinical trial. | ⊗ **Group sequential**<br>○ Sample size reestimation<br>○ Patient population<br>○ Treatment arm selection<br>○ Patient allocation: CAR<br>○ Patient allocation: RAR<br>○ Endpoint selection<br>○ Multi-arm, Multi-stage | No details provided. |
| RHAPSODY-2<br>2022<br>USA and non-USA sites (undefined)<br>High income<br>Investigator-initiated, industry-supported | ⊗ Registration<br>○ Protocol<br>○ Results | Acute<br>Pharmacological<br>Hyperacute<br>90d post<br>NCT05484154 | Multicentre, randomized, placebo-controlled, double-blind, | ⊗ **Group sequential**<br>○ Sample size reestimation<br>○ Patient population<br>○ Treatment arm selection<br>○ Patient allocation: CAR<br>○ Patient allocation: RAR<br>○ Endpoint selection<br>○ Multi-arm, Multi-stage | The study will be conducted in two phases. |
| UPLIFT<br>2022<br>Australia | ⊗ Registration<br>○ Protocol<br>○ Results | Rehabilitation<br>Behavioural<br>Late subacute-chronic | Umbrella design with four independent single | ⊗ **Group sequential**<br>○ Sample size reestimation<br>○ Patient population | Within this umbrella trial, simultaneous Bayesian Optimal phase IIa (BOP) studies will be run to evaluate individual UPLIFT interventions. For each |

| | | | | | |
|---|---|---|---|---|---|
| High income<br>Investigator-initiated | | 4w post intervention<br>ACTRN12622000373774 | arm Bayesian Optimal Phase IIa trials. | ○ Treatment arm selection<br>○ Patient allocation: CAR<br>○ Patient allocation: RAR<br>○ Endpoint selection<br>○ Multi-arm, Multi-stage | intervention, the rate of promising response (composite good outcome) will be monitored at equally spaced, predefined stopping points. Too few promising responses will mean an individual UPLIFT intervention is stopped and future trial participants are directed into other intervention studies under investigation. |

Notes: [a]Income status from The World Bank https://datatopics.worldbank.org/world-development-indicators/the-world-by-income-and-region.html defined as either high-income, upper-middle income, lower-middle income, low-income. [b]Support defined via trial sponsorship and funding definitions across sources as industry-sponsored, investigator-initiated industry-supported, investigator-initiated. Industry supported means that there was substantive involvement such as supply of drug or device, while the funding may have originated from a government body (such as NIH, NHMRC). [c]Population defined as acute, rehabilitation, or prevention. [d]Treatment type defined as pharmacological, device or behavioural. [e]Timepoint of enrolment timepoints defined using the Stroke Recovery and Rehabilitation Roundtable recommendations[36] of 0-24h hyperacute, >24h-7d acute, >7-90d early subacute, >90-180d late subacute and beyond 180d chronic. [f]Registration NCT=clinicaltrials.gov, ACTRN=Australian and New Zealand Clinical Trial Registry. [g]Adaptations to patient allocation CAR = covariate-adaptive randomisation. Adaptations to patient allocation RAR = response-adaptive randomisation. AIS=acute ischaemic stroke. DSMB=data and safety monitoring board. ET=endovascular thrombectomy. ICH=intracerebral haemorrhage. LVO=large vessel occlusion. mRS=modified Rankin Scale. TIA=transient ischaemic attack. MT=mechanical thrombectomy. y=years. m=months. w=weeks. d=days. h=hours. min=minutes. *Results known to have been presented at international meetings but publication of results not publically available at time of submission.
